# Deep Learning Models Can Predict Violence and Threats Against Healthcare Providers Using Clinical Notes

**DOI:** 10.1101/2024.05.27.24308001

**Authors:** Nicholas J Dobbins, Jacqueline Chipkin, Tim Byrne, Omar Ghabra, Julia Siar, Mitchell Sauder, R Michael Huijon, Taylor M Black

## Abstract

Violence, verbal abuse, threats, and sexual harassment of healthcare providers by patients is a major challenge for healthcare organizations around the world, contributing to staff turnover, distress, absenteeism, and reduced job satisfaction and overall mental and physical health. To enable interventions prior to possible violent episodes, we trained two deep learning models to predict violence against healthcare workers 3 days prior to violent events for case and control patients. The first model is a document classification model using clinical notes, and the second a baseline regression model using largely structured data. Our document classification model achieved an F1 score of 0.75 while our model using structured data achieved an F1 of 0.72, both exceeding predictive performance of a psychiatry team who reviewed the same documents (0.5 F1). To aid in explainability and understanding of risk factors for violent events, we additionally trained a named entity recognition classifier on annotations of the same corpus, which achieved an overall F1 of 0.7. This study demonstrates the first deep learning model capable of predicting violent events within healthcare settings using clinical notes, surpassing the first published baseline of human experts. We anticipate our methods can be generalized and extended to enable intervention at other hospital systems.

## Introduction

Workplace violence and harassment toward healthcare providers is a major challenge for healthcare organizations around the world [1]. Evidence suggests that up to 38% of healthcare providers suffer violence in the workplace at some point in their careers, and incidents of physical intimidation, threats, verbal abuse, and sexual harassment are also common [2]. Research also indicate that the frequency of violent events and harassment of healthcare providers has continued to increase since the COVID-19 pandemic [3–5], exacerbating provider burnout and leading to increased stress, staff turnover, absenteeism, and reduced job satisfaction and taking an overall toll on mental and physical health [1]. For medical inpatients, psychiatrists may be consulted to diagnose, treat, or de-escalate when staff perceive possible risk, but staff-initiated requests for consultation are often too late or never begun. At the same time, recent advances in machine learning and natural language processing (NLP) have demonstrated significant potential in training and fine-tuning sophisticated screening models capable of matching or even surpassing the capabilities of human experts [6–9], and often doing so automatically, far faster, and at greater potential scale. We hypothesized that NLP models trained on corpora of clinical notes preceding past violent events could be utilized to prospectively determine patients and staff at risk of violence, enabling intervention and prevention.

Predicting violence against healthcare workers is challenging for a number of reasons. Actuarial models of suicidal or recidivistic criminal violence are notoriously poor, lacking dynamic input. Real-time data allows for much better violence prediction, but requires time for universal screening to capture dynamic risks and complex interplays among patient-level, ward-level, and hospital/social level factors. Patients’ histories, illnesses, perceptions and behavioral symptoms clearly define risks, as captured in screening measures such as the ABRAT [10]. Violence is more likely against individual healthcare workers with less experience, who are themselves insecure [11] and anxious [12], often in conditions of overwhelming traumatic stress. Violence is more prevalent in clinical environments where staff are too busy or too disorganized to respond to these acute needs [13], reflecting institutional and social inequities and management decisions. The concentration of patients with high static risk for violence in under-resourced clinical environments staffed by inexperienced and traumatized healthcare workers is a recipe for driving burnout and turnover [14], compounding the deficit in experienced and behaviorally skillful staff. The use of force (whether through security staff, physical restraints, or chemical sedation) is typically initiated by frontline healthcare workers who may have the least skills and experience, and is influenced by dynamic processes of bias and fear perception which are strongly determined by patients’ race, poverty, and psychiatric labeling. Additionally, many incidents of violence or intimidation go unreported to hospital administrators [15], limiting quality improvement and safety initiatives [16].

For violence prevention interventions to be clinically valuable, they must encompass this complexity. Predictive tools must provide enough time for a clinical response. Clinical services must be tailored to the different phenotypes of patient-level violent behavior and the needs of frontline healthcare workers at risk. These interventions must also be considered at the level of management units and healthcare systems in order to address training, retention, and resilience of effective healthcare workers, as well as the inequities and biases that can exacerbate dynamic risks for violence in ill and injured patients. A few published studies have attempted to develop models capable of predicting violence against healthcare providers. Airakinsen *et al*. surveyed public sector workers in Finland, including nurses, to create logistic regression models for predicting violence or threats for a given worker over time [17]. Kowalenko *et al*. followed over 200 healthcare providers in an emergency department over 9 months and developed a linear regression model for prediction [18]. In both studies, as prediction focused on survey-based education, experience, and demographic characteristics of a given worker, the resulting models were not predictive of violence for a given patient or moment in time, limiting their utility for prospective interventions. Lata and Navel used logistic regression and other methods for predicting non-violent workplace behavior, though this was not specific to healthcare and limited to incidents of non-violent threats. Lee et al. used structured data from ED visits from a tertiary hospital in the Republic of Korea and achieved an F1 score in using a random forest model of 0.84 [19]. However, the study was conducted in a hospital setting under uniquely severe overcrowding and predicted violence likelihood across the entire time-span of an encounter (potentially days or weeks), rather than a specific moment in time. Locality-specific EHR workflows, data quality and cultural factors may further limit generalizability.

To the best of our knowledge, no published studies have attempted to use the contents of clinical notes for predicting violence in a healthcare setting or predicted violence for a specific moment in time. While the unstructured, free-text nature of clinical notes make them somewhat challenging for analysis, narratives within clinical notes are also an incredibly rich source of psychosocial information, containing nuance and detail often absent from corresponding structured data [20, 21]. We hypothesized that training a deep learning model using clinical notes could yield novel insights and high predictive performance.

### Key Contributions

This study contributes the following:

1. **A novel benchmark and clinical note corpus of predictions of forthcoming violence** by human experts trained in psychiatry.
2. **A high-quality double annotation of the same corpus for violence risk factors**, such as aggressive behavior, cognition, psychosis, and substance abuse.
3. **A baseline regression model using largely structured data** which achieves an F1 of 0.72.
4. **A named entity recognition (NER) model for predicting violence risk factors** in clinical notes which achieves an overall 0.7 F1.
5. **The first document classification model able to predict violence against healthcare workers for a specific patient and moment in time**, with 0.75 F1, exceeding that of our baseline regression model (F1 0.72) and human psychiatry team (F1 0.5).

Figure 1 shows our overall study design.

**Fig. 1.**
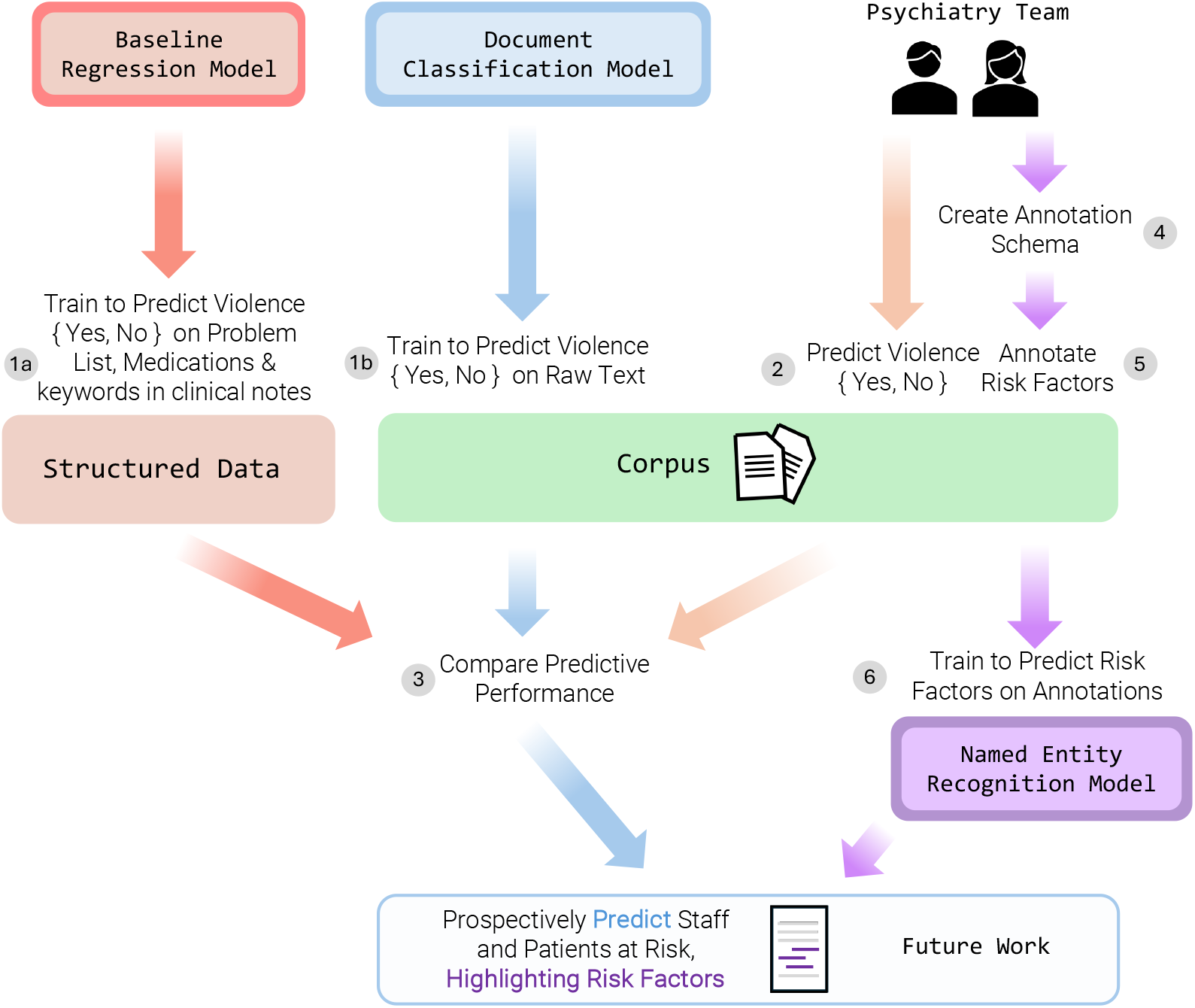
Overall study design. (1) We labeled our corpus with 1 (cases) or 0 (controls) and (1a) trained a regression model using largely structured data and (1b) trained a Clinical-Longformer model for document classification as a regression task. (2) In pairs, our psychiatry team reviewed each document and predicted {Yes, No} that a document preceded a violent event. If a pair disagreed, a third psychiatrist reviewed it as a tie-breaker. (3) We compared the combined predictive performance of the psychiatry team across the entire corpus (Train + Test) to that of the document classification model (Test only). (4) Our psychiatry team leaders developed an annotation schema for risk factors for violence. (5) As the psychiatry team reviewed documents, they additionally annotated risk factors. The annotation schema was iteratively revised based on team feedback. (6) We trained a named entity recognition model for predicting risk factors.

## Methods

### Dataset

With the approval of our hospital leadership, we obtained two datasets of incidents from March 2021 to October 2023 involving hospital security.

1. *Code Grays* are security staff response incidents of patients attempting to leave or resist care when this poses imminent danger.
2. *Staff Patient Safety Net* (PSN) incidents involve violence against hospital staff, often leading to significant staff distress.

The two events often coincide. We first combined, then de-duplicated the two datasets in order to ensure a given incident was recorded only once. Many Code Gray and PSN events are preceded by staff taking precautions when patients appear potentially violent or threatening, often mentioned in clinical notes. As we aimed to limit our predictions to only cases where the event was surprising and unanticipated – and thus prediction beforehand would be of most value – we filtered out any events in which, in clinical notes in the 3 days preceding the timestamp of the event, we found any mention of terms indicative of security monitoring, such as “1:1”, “sitter”, “detained”, “against medical advice”, and so on. This resulted in 280 cases with unique timestamps from 246 unique patients. Using these 280 cases, we extracted clinical notes in the 3 days preceding a given timestamp. As patients often have many clinical documents written, some less potentially useful for our analysis, we limited clinical notes to only those of “H&P” (history and physical) and “Nursing Note” types. H&P notes are typically longer and describe patient history and reason for hospitalization, recent labs and medication orders and so on, and plan for care. Nursing notes tend to be shorter, more frequent updates on patient status. For each of the unique events, we concatenated the notes into a single long document.

### Case-Control Matching

We achieved a 1:1 matching of cases and control patients. To do so, we wrote an algorithm querying our data warehouse to match cases to a control patient of the same biological sex, same age within +/-5 years, admitted in the same 2.5 year time window, and with the highest number of matching ICD-10 diagnosis codes for the encounter, limited to patients with an H&P note written. For example, a male case patient who was 46 years old at the time of the violent event and admitted with diagnosis codes for Altered Mental Status (R41.82), Wheezing (R06.2), and Tachycardia (R00.00), would be matched to the first male control patient aged 41-51 and with the same admitting diagnoses found. If no patient had all 3 diagnosis codes, the first patient with 2 of the 3 would be selected, and so on. This resulted in 280 corresponding control patients. We used the time 3 days after the initial H&P note as an artificial timestamp for controls and similarly created a long document for each. Our combined case + control dataset thus had 560 total documents.

### Risk Factor Annotation Schema and Annotation

Because our long-term goal in future work is to prospectively identify at-risk providers and patients in order to intervene and de-escalate potential violent events, ranking risk as a regression task alone (i.e., predicting a normalized value between 0.0 and 1.0) does not lend sufficient explainability. For example, if a psychiatry team received a daily report of providers and patients at risk but the report showed only a single predicted continuous value alongside each patient (e.g., “John Doe: 0.387”), it would be impossible to understand why a model output such a prediction without actually reviewing a patient’s chart - with the caveat that even then, human understanding and factors influencing model prediction may not align. While possible solutions such as SHAP [22] could be utilized to highlight segments of clinical notes correlated to a given model output, highlights across an entire set of long concatenated notes could be time-consuming to read, when instead a short summary would be preferable. We therefore reasoned that developing an annotated corpus of risk factors for violence from the perspective of psychiatrists could be used to train a NER model and aid in explainability and summary for such a future report. Risk factor identification could thereby guide actionable, targeted interventions such as substance use treatment, suicide precautions, or delirium management.

Our lead psychiatry team members developed an annotation guideline for the following 8 categories:

1. **Aggressive Behavior** - Aggressive actions observed during hospitalization and those performed prior to admission. Observed behaviors were based on the validated Broset Violence Checklist [23], a validated, short-term violence prediction instrument. Previous actions included interpersonal violence perpetration and victimization, such as history of assaults or suicide attempts.
2. **Cognition** - The six neurocognitive functioning domains: 1) memory and learning, 2) language, 3) executive functioning, 4) complex attention, 5) social cognition, and 6) perceptual and motor functioning. In clinical documentation, cognitive impairments are often reported through a patient’s levels of alertness, orientation status, and ability to comprehend medical care and communicate medical decisions.
3. **Mood symptoms** - Based on DSM-5 symptomatology for depressive, anxious, and manic conditions, this category identified disordered alterations in patients’ emotional states during the current hospitalization such as suicidal ideation, hopelessness, rumination, panic and grandiosity.
4. **Psychotic symptoms** - A loss of contact with reality. This category included positive symptoms such as delusions, hallucinations, and disorganized thoughts and behaviors, and negative symptoms such as emotional blunting, avolition, and poverty of thought.
5. **Acute substance use** - The recent recreational use of mood-altering substances, both legal (such as nicotine, alcohol, and cannabis) and illicit (such as opioids and stimulants). Recent use prior to or during admission could be self-reported or referenced through toxicology results. In addition, this category included signs and symptoms of active substance intoxication, withdrawal, and craving.
6. **Unmet needs / interpersonal conflict** - Patient-reported dissatisfactions with care. This included concrete complaints such as poorly controlled pain, interrupted sleep, rescheduled procedures, and premature or delayed discharge. It also included abstract patient perceptions of mistreatment by medical providers, such as feeling ignored or judged.
7. **Noncompliance** - Patient refusal to participate in medically necessary care. This included refusing to take scheduled medications, follow physical restrictions, and participate in clinical interviews, exams and diagnostic interventions such as lab draws and imaging studies. It also included purposeful removal of medical equipment such as braces or bandage drains.
8. **High care utilization** - A disproportionate burden on the healthcare system due to elevated resource use. This included references to bounce-back medical admissions, frequent emergency room visits, past psychiatric hospitalizations (for either voluntary and involuntary treatment), past incarcerations, and past or current treatment with community mental health organizations.

For our 560 total documents, we enlisted the help of six total annotators of various levels of experience in psychiatry: two attending psychiatrists, three psychiatry residents, and one medical student entering psychiatry residency. In addition to annotating risk factors, we also utilized our psychiatry team to establish a baseline of how well human experts can predict an upcoming violent event by reading clinical notes. To do so, at the end of each document we added the text *<< CODE GRAY OR PSN WILL OCCUR >>*, which annotators were instructed to annotate as Yes or No based on their experience and intuition. In order to ensure a high quality of annotation, we first trained all annotators on the same 20 randomly selected documents, then copied all annotation variations for each training document into a “differential” file from which the annotation team used to reconcile differences. We then double-annotated the remaining documents by pairing annotators and randomly assigning 135 documents to each group, split into batches. After each round of annotation was complete, we generated differential files for each pair and completed reconciliation. Two of the annotators were paired twice, and thus annotated approximately twice the number of documents as others. If a pair differed in their Yes/No prediction of violence in a given reconciled document, a 3rd “tie-breaker” psychiatrist from our annotation leaders reviewed the document and determined a final Yes/No prediction.

### Baseline Regression Model

In addition to our human psychiatry group, we also aimed to create an additional baseline regression model using largely structured data elements. We based the inputs to this model on the MEND Screening Model at the University of Pennsylvania [24], using patient age at prediction, psychiatric diagnoses associated with medication orders, presence of active mood disorders and anxiety-related diagnoses on the problem list or past billing diagnosis codes, whether antidepressants were administered in the encounter, and prior ED visits with psychiatric complaint within past two years. Additionally, we explored the inclusion of keyword items in preceding clinical notes relating to anxiety, abuse, psychiatry, depression, withdrawal, ativan, alprazolam, and intravenous drug use.

### Evaluation

At a high level, our evaluation sought to answer the following questions:

**1. How well can a document classification model predict violence? How does this compare to human experts or models using structured data?** Alongside our human and structured data baseline, we aimed to create a deep learning model capable of matching or surpassing human experts in prediction, trained on an unannotated raw text dataset. As documents in our dataset tended to be long, we fine-tuned the Clinical-Longformer base model [25] for document classification after randomly splitting our dataset into an 80/20 split of train and test documents. In cases where a given document exceeded the length of the context window allowed by Clinical-Longformer or our GPU memory constraints, after some experimentation, we found that cropping the document by taking only the beginning and ending (up to 50% of characters allowed in the context window on each side), effectively removing the middle of the document, to work reasonably well.

**2. How well can an NER model predict risk factors?** Using the risk factor annotations by our psychiatry team, we trained a NER model to predict an output label for each token in a given document. In order to evaluate every word in a given document, we used a moving-window strategy, splitting each document based on the maximum allowed tokens for a given model’s context window, then evaluating each window of text using the Bio ClinicalBERTbase model [26].

We use the F1 score as our primary evaluation metric in both tasks, where F1 = 2 * (precision * recall) / (precision + recall).

All experiments were approved by our institutional review board (IRB #00018889).

## Results

Results of our first experiment are shown in Table 1. After annotations and tie-breaks of predictions were completed, the psychiatry team had an overall precision of 0.62, recall of 0.41, and F1 of 0.5, with the top individual at 0.62, 0.53, and 0.57 respectively. Some psychiatry team members annotated more documents than others based on availability. Our Clinical-Longformer model, adapted for a regression task, outputs a value between 0 and 1 which we normalize and compare to an optimized threshold for prediction. Table 1 shows results for 2 thresholds, one optimized for recall (0.75) and one for precision (0.78), both of which achieve an F1 of 0.75, exceeding that of our human psychiatrist team. Our baseline regression model also achieved reasonably good performance and surpassed the psychiatrist team with a somewhat lower F1 of 0.72.

**Table 1.**
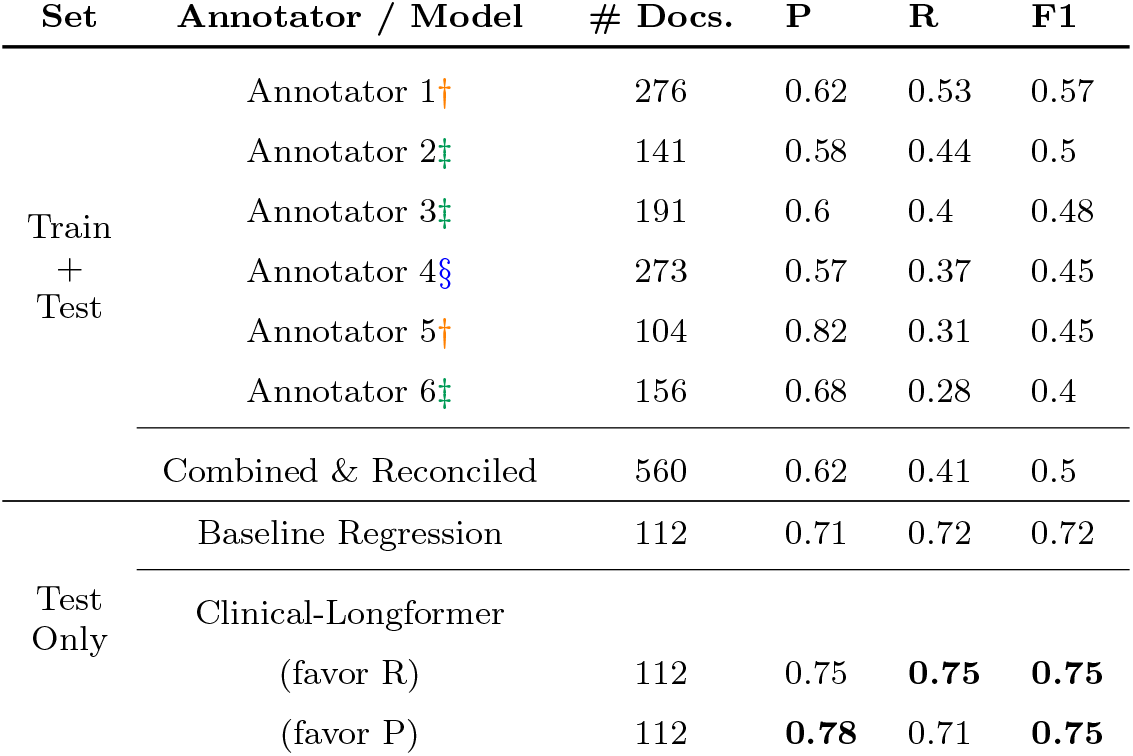
Results of our violence prediction evaluation comparing humans trained in psychiatry compared to our baseline regression and fine-tuned Clinical-Longformer model. The human annotators are paired, and thus annotated documents are counted twice, but only once per annotator. Due to differences in availability for annotation, certain annotators annotated more documents than others. The annotators include Attending Psychiatrists†, Psychiatry Residents‡, and a Medical Student entering Psychiatry Residency§.

We focused subsequent analysis on our Clinical-Longformer model as it achieved the highest performance. Figure 2 shows the Receiver Operating Characteristic (ROC) curve using a threshold at 0.18, favoring precision, as well as distribution of test set patients by ethnic heritage. Table 2 similarly shows P/R/F1 and accuracy for test set patients by ethnic heritage.

**Table 2.** Test set document classification evaluation results by ethnic heritage. One test patient had unknown ethnic heritage and was excluded.

| Ethnic Identification | # Patients | P | R | F1 | Acc. |
| --- | --- | --- | --- | --- | --- |
| American Indian or Native Alaskan | 5 | 1.0 | 0.75 | 0.86 | 0.8 |
| Asian | 6 | 0.0 | 0.0 | 0.0 | 0.5 |
| Black or AfricanAmerican | 15 | 1.0 | 0.7 | 0.82 | 0.8 |
| Multiple | 3 | 0.5 | 0.5 | 0.5 | 0.33 |
| Native Hawaiian or Pacific Islander | 1 | 0.0 | 0.0 | 0.0 | 0.0 |
| White | 81 | 0.76 | 0.79 | 0.77 | 0.78 |

**Fig. 2.**
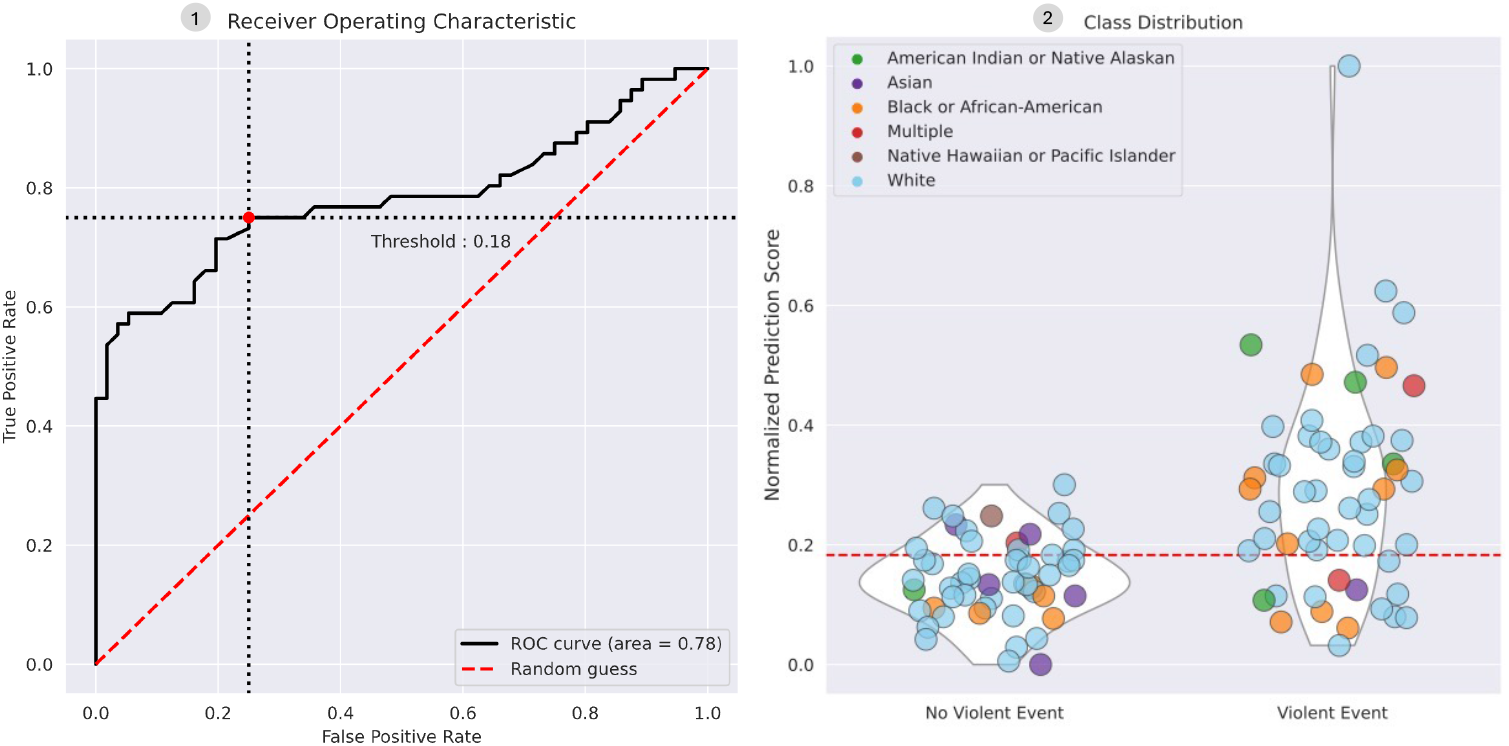
Left - Receiver Operating Characteristic (ROC) curve for the Clinical-Longformer model with threshold at 0.18, which favors precision. Right - violin and bubble plots of test set patients, with Y axis showing normalized prediction scores. Bubbles are color coded by patient ethnic identification.

We additionally conducted analysis using SHAP values to better understand phrases and documents which correlated with higher and lower normalized prediction scores. Figure 3 shows the extracted mean SHAP values by tokens and phrases within the test set. Many, though not all, strongly correlated phrases are reasonably intuitive, with phrases correlated toward non-violent documents often reflecting reassuring nursing language (e.g., “pleasant and cooperative”), while phrases such as “PTSD” or relating to methamphetamine use correlated with violence. Figure 4 shows an example test set document with risk factor annotations by a psychiatrist on the left and SHAP values from the Clinical-Longformer model for the same document on the right. Remarkably, we found SHAP values often overlap with human-annotated risk factors, despite training the model on only raw, unannotated document text and violence outcomes.

**Fig. 3.**
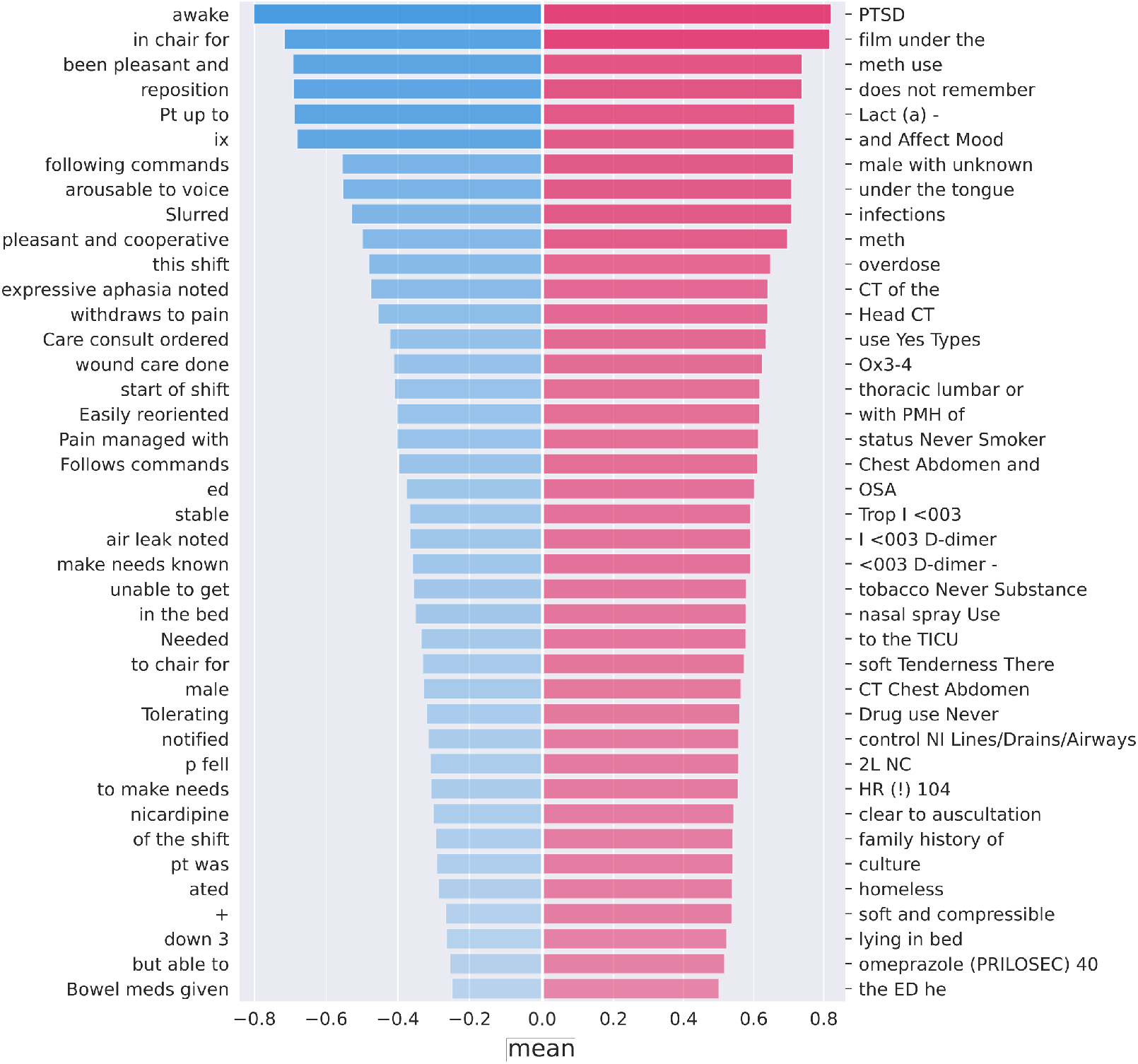
The top 40 phrases we found to influence model predictions toward violent events (red) and not (blue), as measured by mean SHAP values across the test set.

**Fig. 4.**
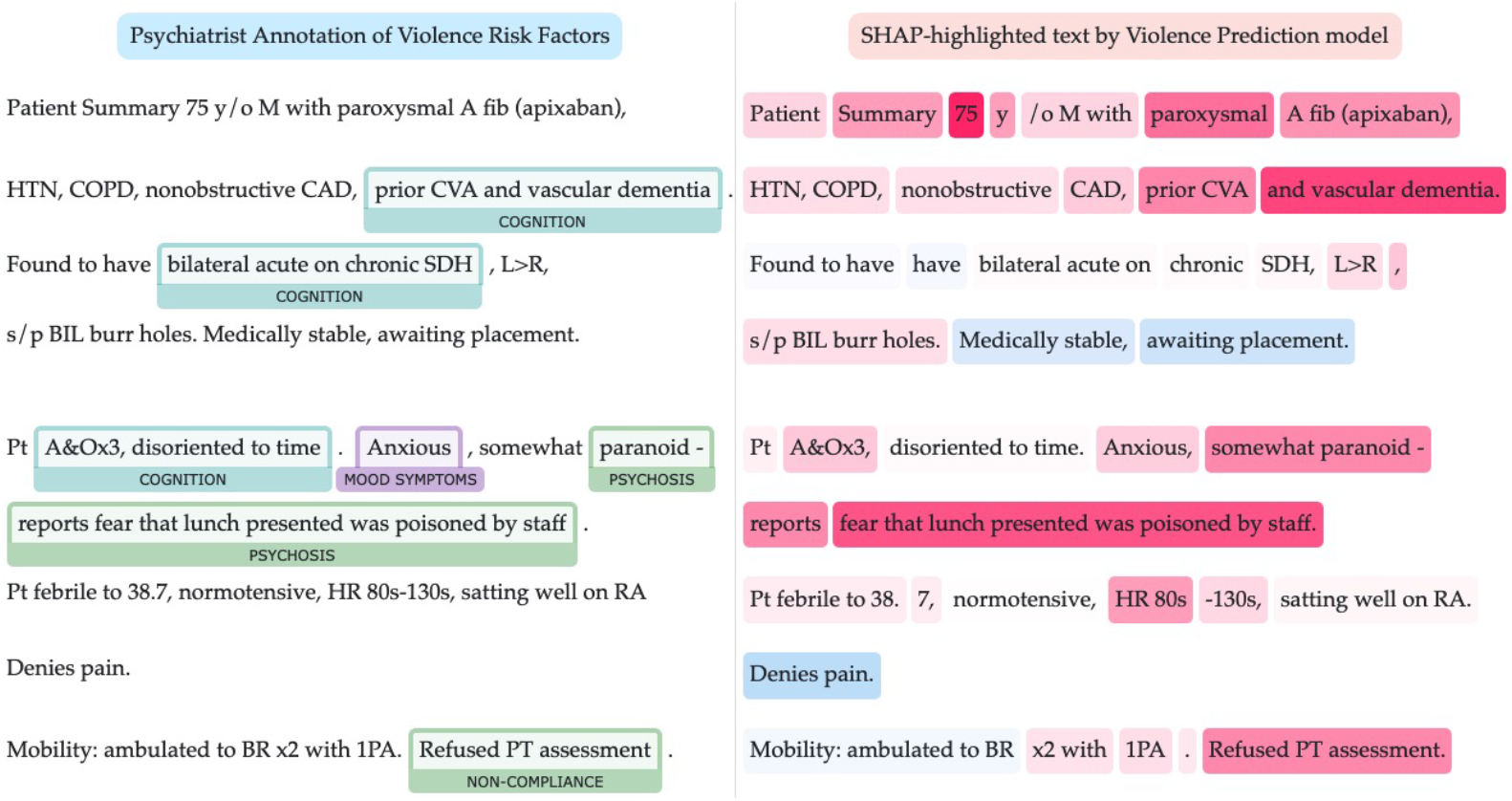
Side-by-side comparison of a psychiatrist annotation of risk factors for violence or threats compared to token-level SHAP highlights correlated to the prediction from our fine-tuned document classification model, abbreviated and edited for clarity. Correlation does not imply causation, but the automatically learned phrases strongly predictive of Code Gray or PSN by the model (right) are in certain areas strikingly similar to psychiatrist annotations (left), despite being trained on raw clinical note text rather than annotations. This example has been abbreviated and modified to prevent patient re-identification.

Results of our second experiment for named entity recognition are shown in Table 3. We calculated both a *strict* scoring strategy, requiring exact matching of annotated and predicted token indices and category, as well as a *relaxed* scoring strategy, which requires that categories match and 50% or more of gold tokens be overlapped with predicted tokens. Our strict scoring strategy resulted in an overall F1 score of 0.65 and relaxed an F1 of 0.7. As we envision future prospective reports highlighting risk factors to include surrounding text context (e.g., the full sentence where a given risk factor was predicted), we find a relaxed scoring strategy to be a reasonable measure of model performance. Among risk factor categories, Cognition showed the highest F1 (strict: 0.71, relaxed: 0.75) while unmet needs showed the lowest (strict: 0.44, relaxed: 0.48).

**Table 3.** Results of our named entity evaluation using Bio ClinicalBERT. The **Strict** scoring requires exact matching of start and end character indices and category, while **Relaxed** scoring requires category matching and half or more of gold and predicted tokens to overlap.

| Category | # In Test Set | Scoring Strategy |  |  |  |  |  |
| --- | --- | --- | --- | --- | --- | --- | --- |
|  |  | Strict |  |  | Relaxed |  |  |
|  |  | P | R | F1 | P | R | F1 |
| Acute Substance Use | 141 | 0.67 | 0.62 | 0.64 | 0.73 | 0.66 | 0.69 |
| Aggressive Behavior | 263 | 0.63 | 0.71 | 0.67 | 0.67 | 0.77 | 0.72 |
| Cognition | 627 | 0.69 | 0.73 | 0.71 | 0.72 | 0.78 | 0.75 |
| High Utilization or Incarceration | 92 | 0.69 | 0.66 | 0.67 | 0.72 | 0.69 | 0.7 |
| Mood Symptoms | 139 | 0.62 | 0.66 | 0.64 | 0.66 | 0.71 | 0.68 |
| Noncompliance | 228 | 0.64 | 0.65 | 0.64 | 0.69 | 0.7 | 0.69 |
| Psychosis | 66 | 0.63 | 0.73 | 0.68 | 0.67 | 0.77 | 0.72 |
| Unmet Needs or Conflict | 175 | 0.43 | 0.45 | 0.44 | 0.47 | 0.5 | 0.48 |
| Total (micro avg.) | 1463 | 0.64 | 0.67 | 0.65 | 0.68 | 0.72 | 0.7 |

## Discussion

Even for trained human experts with extensive knowledge of a local patient population and documented risk factors, prediction of violence remains challenging. However, our experiments demonstrate that deep learning models can predict forthcoming violent events toward healthcare providers with reasonably strong recall and precision. Our Clinical-Longformer model performed somewhat better than our baseline regression model using mostly structured data, but both achieved an F1 of over 0.7, compared to 0.5 for our psychiatrist team.

Our risk factor prediction NER model trained on psychiatrist annotations also serves as a novel means of providing visual context alongside a numerical risk score. While our NER model may not capture the same information presented in a SHAP visualization, we believe it presents much of the same and most salient information from the perspective of psychiatrists and also lends itself more easily toward summary and clarity. We hope the outputs from our two models together therefore may enable a future psychiatry team prospectively viewing risk scores to quickly understand risk factor context and make appropriate informed decisions in terms of prioritizing interventions.

As we intend to prospectively deploy our models to detect forthcoming violence and enable intervention and de-escalation, we also considered concerns around model bias based on patients’ ethnic heritage and racial identity. As can be seen in Table 2, using the threshold optimized for precision on the entire test set of 112 documents, the model performed reasonably well for patients identifying as American Indian or Native Alaskan (F1 0.86), Black or African-American (F1 0.82) and White (F1 0.77), but poorly for Asian and Native Hawaiian patients (F1 0.0 for both), though given the relatively small sample size further study on a larger set of patients is needed.

### Limitations

This study had a number of limitations. First, we used clinical notes and data from only one institution and a relatively small window of time (2.5 years). External validation is needed to test if our results will generalize to other hospitals. In the future, we intend to work with colleagues at other safety net hospital systems and utilize techniques such as federated learning [27] to leverage multiple sites’ data and improve models without sharing data directly with one another. We also explored and compared SHAP values and token highlights correlated to model predictions, though correlation does not imply causation. Our comparison to psychiatrist-annotated risk factors, while visually similar in many cases, nonetheless requires further study and empirical analysis to better understand how model learnings relate to psychiatric training and evaluation methods.

### Future Work

We intend to explore methods for further improving predict performance, including exploration of models which combine structured and clinical note data as inputs, as well as preprocessing and integrating risk factor predictions into the document classification inputs to evaluate whether that improves our models as well. Additionally, we intend to explore the piloting of our models for prospective violence prediction and possible intervention, as well as robust evaluation of such predictions for bias and possible harm before deployment.

## Data Availability

Due to the sensitive nature of data used in this study, we are currently unable to make the data used available upon request.

## Code Availability

Code used for data preparation, preprocessing, training, and evaluation is available at https://github.com/ndobb/codegray^1^.

## Footnotes

1 Code will be made available upon article acceptance

